# Aphasia in Bilinguals

**DOI:** 10.1101/2020.04.22.20075432

**Authors:** Alfredo Ardila, Durjoy Lahiri

## Abstract

**Background:** Little is known about the influence of bilingualism in aphasia.

**Aims:** To analyze the effect of bilingualism in a group of patients with aphasia.

**Methods:** Data from155 monolingual and 53 bilingual aphasia patients were analyzed.

**Results:** It was found that aphasia was significantly less severe in bilinguals when compared to monolinguals (p=0.023)

**Conclusion:** Bilingualism represents a protecting factor in aphasia. Aphasia is less severe and consequently, recovery is expected to be better in bilingual patients.

## Introduction

It has been reported that bilingualism represents a type of cognitive reserve (e.g., Bialystok, Craik, Klein, & Viswanathan, 2004; Gold, Johnson, & Powell, 2013) and delays the onset of dementia (e.g., Alladi, et al., 2013; Bialystok, Craik, & Freedman, 2007). Bilingualism has also been associated with increase of some specific cognitive abilities, including but not limited to executive functions (Bialystok, 2011, 2015), spatial tasks (Greenberg, Bellana & Bialystok, 2013; McLeay, 2003), and working memory (Luo, Craik, Moreno, & Bialystok, 2013). This perspective, however, remains controversial and sometimes no bilingual advantage on cognition is reported (e.g., Dick et al., 2019). Calvo, García, Manoiloff & Ibáñez (2016) in a review paper found that only that around 60-70% of published data have found a cognitive reserve effect of bilingualism on cognitive decline. Recently Lehtonen et al. (2018) compared the bilinguals’ and monolinguals’ performance in six executive domains based on 152 studies on adults. Their analyses revealed a very small bilingual advantage in inhibition, shifting, and working memory, but not in monitoring or attention. The authors concluded that available evidence does not support the widely held notion that bilingualism is associated with benefits in cognitive control functions in adults.

Bilingualism affects the cognitive outcome after stroke. Alladi et al. (2016) studied 608 patients with ischemic stroke and analyzed the effect of bilingualism on post-stroke cognitive impairment. No differences in frequency of aphasia were observed: 11.8% of monolingual participants presented aphasia, compared to 10.5% of the bilingual patients (p<0.354). However, twice the number of bilingual patients had normal cognition compared to monolinguals (40.5% and 19.6% respectively) (p<0.0001). The reverse pattern was observed when cognitive impairment was analyzed: in 77.7% of monolinguals and 49.0% of bilinguals cognitive impairment was documented (p<0.0009). The authors concluded that bilingualism is associated with a better cognitive profile after stroke, probably due to an increased cognitive reserve.

Penn, Frankel, Watermeyer, and Russell (2010) suggested that bilingualism could affect the aphasia characteristics. They found that bilingual individuals with aphasia presented superior conversational skills; these increased skills were correlated with better executive functions compared to monolinguals, and hence, their better performance in expressive language was associated with a higher executive control. Hope and colleagues (2015), however, reported that bilingual immigrants, who were non-native English speakers with aphasia performed worse in several language tests administered both in L1 (native language) and L2 (English) compared to monolingual native English-speaking aphasia patients. The authors suggested that in this specific bilingual sample, poor premorbid language proficiency can be assumed in comparison with the monolingual control patients; they further hypothesized that bilinguals may be more sensitive to brain pathological conditions. Moreover, Faroqi-Shah, Sampson, Pranger, & Baughman (2018) studied 38 individuals with aphasia. They were administered a task measuring cognitive control (Stroop color-word task) and two word production tasks (picture naming and category fluency). The authors found lower cognitive control in the aphasia group relative to age-matched neurologically healthy adults. However, a bilingual advantage in cognitive control was found in neurologically healthy adults and in one group of bilingual speakers with aphasia, but not the other group.

It has been observed that bilingual individuals with aphasia show more efficient alerting skills than monolinguals with aphasia (Dash, Masson-Trottier, & Ansaldo, 2020). Bilinguals with aphasia can also present an advantage in inhibitory control tasks (Faroqi-Shah, Sampson, Pranger & Baughman, 2016). Differences in language and bilingual control have been documented in in bilingual individual presenting aphasia (Gray & Kiran, 2015, 2019).

Information about the influence of bilingualism on aphasia severity is limited, regardless the abundant literature about the characteristics and recovery patterns of both languages in bilingual aphasia (e.g., Fabbro, 2001; Faroqi-Shah, Frymark, Mullen & Wang, 2010; Lorenzen& Murray, 2008; Paradis & Libben, 2014). Two papers dealt directly with the issue of aphasia severity in bilinguals. Paplikar et al. (2019) studied 38 bilingual and 27 monolingual aphasia patients. In all the cases, aphasia was due to stroke. Language was evaluated at least three months after the stroke(mean 11.5 months). The Addenbrooke’s CognitiveExamination – Revised (ACE-R) was used for the diagnosis of aphasia. Monolinguals’ and bilinguals’ scores were compared after controlling for confoundingvariablesincluding: age, gender, education, occupation,medical, and stroke characteristics. Aphasia severity was significantly higher in monolinguals compared to bilinguals(7.0 vs. 14.4, maximum score 40; p = 0.008).Bilinguals also had a better performance in the attention, memory, and visuospatial domains of ACE-R. A univariate general linear model analysis found that bilingualism was significantly associated with higher language scores in the ACE-R after adjusting for potentially confounding variables.It was concluded that although bilingual speakers have a similar risk of developing aphasia after stroke, their aphasia is likely to be less severe.

More recently Dekhtyar, Kiran, & Gray (2020) studied examined in 13 Spanish-English bilingual healthy adults (BHA), English monolingual healthy adults (MHA), Spanish-English bilingual adults with aphasia (BAA) and 18 English monolingual adults with aphasia (MAA). In the last two groups participants were matched by age, education, language impairment, and non-verbal executive functions. The authors report that no evidence of bilingual cognitive control advantage on reaction times in healthy adult groups; however, BAA were faster than MAA, suggesting that bilingualism may contribute to cognitive reserve in adults with aphasia. The authors concluded that bilingualism may represent a protection factor after an acquired brain pathology.

The aim of current study was to analyze the effect of bilingualism on aphasia severity in a sample of bilingual patients. Their results were compared with a groups of monolingual aphasia patients.

## Methods

Data from155 monolingual and 53 bilingual aphasia patients in a hospital in Kolkata (formerly Calcutta), India, were analyzed.

### The Kolkata Aphasia Study

An observational study in the stroke section of a neurology center of Kolkata, India, was developed. Participants were individuals with first ever strokes. A two-year period (between 2016 and 2018) was used. Bengali is the official language of Kolkata (East India). For all the patients Bengali was the mother tongue, but some of them could also speak English or other Indian national language. Approval from the Institutional Ethics Committee was obtained before beginning the study. The general results of Kolkata Aphasia Study are presented in several papers (Lahiri et al., 2019a, 2019b, 2019c, 2020)

### Participants

Consecutive patients with first ever acute stroke were recruited for this study. We used the following inclusion criteria: (1) alert at the moment of language assessment; (2) only literate adults over 18 years were included; (3) capable to communicate in Bengali language. Exclusion criteria were: (1) general cognitive defects affecting language assessment;(2) dementia documented or suspected; (3) pre-morbid psychiatric disorders hindering communication;(4) abuse of alcohol or drugs; (5) aphasia as consequence of vascular intra-cranial space-occupying lesion

Bilingualism was understood as the ability to communicate in two or more languages (Mohanty, 1994). This information was obtained from the patient and/or the family. In India, there is a high number of spoken languages, and bilingualism is relatively frequent.

### Aphasia Assessment

The Bengali version of Western Aphasia Battery (BWAB) (Keshree, Kumar, Basu, Chakrabarty, & Kishore, 2013) was used for aphasia assessment. In this test battery, an Aphasia Quotient (AQ) can be calculated; aphasia diagnosis is given when this quotient is lower than 93.8. BWAB was administered between the 3^rd^ and 7^th^day following stroke. Magnitude of aphasia severity was determined using the AQ score. Additionally, patients were divided into two groups using a cut- off point of 50: non-severe aphasia (AQ 50 or more) and severe aphasia (AQ less than 50). In severe aphasia lesion mean volume was 18.08 mm^3^ (SD=1.54) whereas in non-severe aphasia it was 6.9 mm^3^ (SD=4.05).

Table 1 presents the demographic variables of the sample and the general results

**Table 1.** Demographic variables of the sample (adapted from Lahiri et al., 2020)

| **Variables** | **Non-Severe**<br>**(n=40)** | **Severe**<br>**(n=168)** |
| --- | --- | --- |
| Age (years) | 51.1(SD=11.79) | 52.44(SD=1.07) |
| Male/Female | 31/9 | 113/55 |
| Education (years) | 8.65(SD=5.06) | 7.59(SD=4.85) |
| Monolingual/Bilingual | 23/17 | 132/36 |

### Statistical Analysis

Two different types of statistical analyses were performed.

1. Initially, bilingual and monolingual participants were compared in three demographic variables (gender, age, and education) as well as in the degree of aphasia severity. Chi-square tests were used for gender and aphasia severity (Non-severe/Severe); t-tests were used to compare age and education.
2. An analysis of covariance (ANCOVA) was developed to find the significance of bilingualism in aphasia severity using Education and Lesion Volume as co-variants. AQ score was furthermore used as a measure of aphasia severity, and lesion extension in mm^3^ was used as a measure of lesion volume.

## Results

Table 2 presents the differences between monolingual and bilingual participants in three demographic variables (gender, age, and education) as well as in aphasia severity. No gender or age effect was found. Interestingly, there was an uneven gender distribution (144 men and 64 women) as well as a low mean age in both groups (52.82 and 50.34 years). Differences in education and aphasia severity between both groups, however, were highly significant. The educational level of the bilingual patients was two and half higher than the education level of the monolingual participants, suggesting that the second language may be acquired at school or that bilingual participants had a higher socioeconomic level than the monolingual participants.

**Table 2.** Differences between monolingual and bilingual participants in three demographic variables and in aphasia severity.

|  | **Monolinguals** | **Bilinguals** | **p** |
| --- | --- | --- | --- |
| Gender (M/F) | 110/45 | 34/19 | 0.224<sup>1</sup> |
| Age (mean(SD)) | 52.82(10.70) | 50.34(11.61) | 0.413<sup>2</sup> |
| Education (mean(SD)) | 5.61(2.91) | 14.19(3.74) | 0.001<sup>2</sup> |
| Non-severe/Severe | 23/132 | 17/36 | 0.007<sup>1</sup> |

<sup>1</sup>Chi-square; <sup>2</sup>t-test

Table 2 presents the differences between monolingual and bilingual participants in three demographic variables and in aphasia severity.

To control the educational effect on the aphasia severity, an ANCOVA entering education was co-variant, was developed. Besides, considering that theoretically the major factor affecting aphasia severity is lesion volume, which could potentially be different in monolinguals and bilinguals, lesion volume was also included as co-variant. Table 3 presents the results of ANCOVA. As anticipated, lesion volume was the major factor determining aphasia severity. The effect of education disappeared in this analyses, and the effect of bilingualism was statistically significant (p=0.023).

**Table 3.**
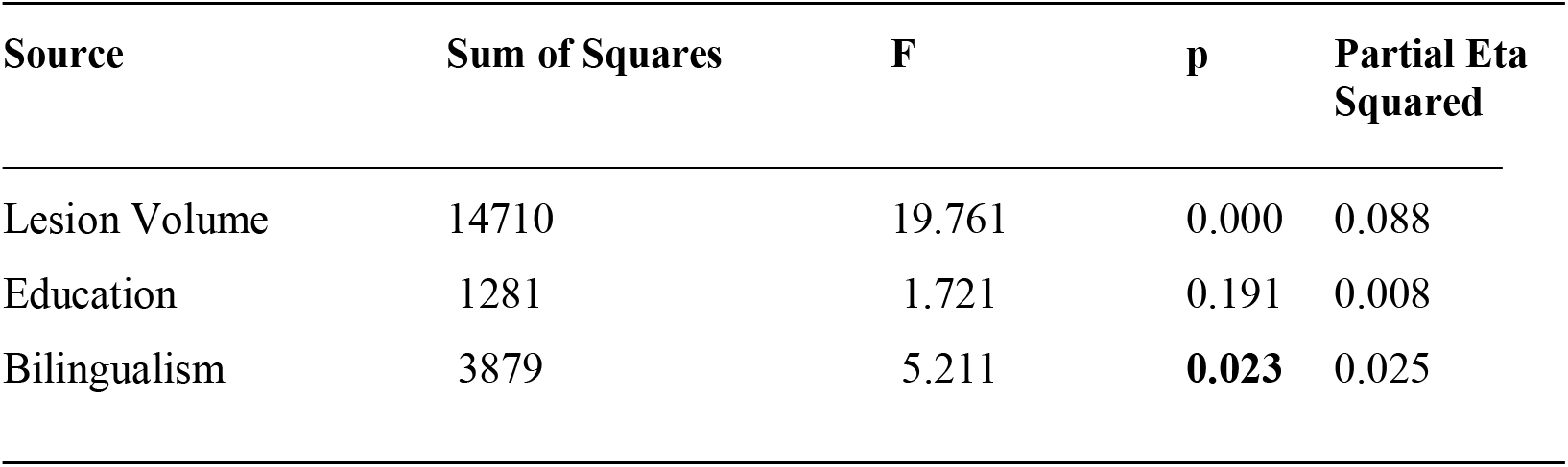
Effect of bilingualism on aphasia severity. ANCOVA using lesion volume and education as co-variants.

| Source | Sum of Squares | F | p | Partial Eta Squared |
| --- | --- | --- | --- | --- |
| Lesion Volume | 14710 | 19.761 | 0.000 | 0.088 |
| Education | 1281 | 1.721 | 0.191 | 0.008 |
| Bilingualism | 3879 | 5.211 | **0.023** | 0.025 |

Finally, to be sure that the difference in aphasia severity between both groups was not a consequence of the lesion size, lesion volumes were compared. Although lesions were slightly larger in monolinguals, difference in volume was not statistically significant (p=-.323)

## Discussion

Few studies have approached the effect of bilingualism on aphasia severity. However, for several years it had been suggested that bilingualism can affect the aphasia characteristics (Penn et al., 2010) and bilinguals could maintain a better performance in at least some verbal abilities.

Paplikar et al. (2019) study represents crucial information in the area; the authors found that language--and also other cognitive abilities--, were better preserved in bilinguals than in monolinguals in cases of stroke.

Nonetheless, the effect of bilingualism in cases of acquired brain pathology does not seem to be limited to the language abilities. Bilingualism has been reported to be associated with less severe cognitive disturbances after stroke (Alladi et al., 2016) and also with better cognitive outcomes (Wood, 2016), suggesting a general protective effect, not restricted to the verbal domain. In Paplikar et al. study (2019), bilinguals had better performance in diverse cognitive areas, including attention, memory, and visuospatial domains; that means, in verbal and also non-verbal abilities.

The current report, corresponding to the Kolkata Aphasia Study, clearly supports that aphasia is significantly less severe in bilinguals, after controlling for potentially confounding variables. As it is obvious, the major factor predicting aphasia severity was lesion volume, but differences in lesion volume were not significant between monolinguals and biinguals. Education initially appeared as a significant factor predicting aphasia severity. But education was highly correlated with bilingualism, and the analysis of covariance demonstrated that the crucial factor was indeed bilingualism, not education.

Current results are congruent with previous research and corroborate that bilingualism is indeed a protecting factor in cases of stroke. That means, bilingualism can be interpreted as a protecting factor in cases of both, progressive (e.g., Alzheimer disease), and also abrupt (stroke) brain pathology, suggesting that indeed bilingualism represents a significant cognitive reserve. Hopefully, toward the future new studies will clarify if this reserve is increased in cases of trilingualism and multilingualism, as found during normal aging (Kavé, Eyal, Shorek, & Cohen-Mansfield, 2008).

Interestingly, gender did not predict aphasia severity (Table 2), but was associated with aphasia probability (144 men and 64 women; that is, men represented 69% of the aphasia sample). This distribution has not been reported in western aphasia samples (e.g., Pedersen, Vinter, & Olsen, 2004), but this distribution is similar to the percentage reported by Paplikar et al. (2019) (75.4% of the patients were male) in another study in India. We do not have any evident explanation for this uneven gender distribution, even though it could be speculated that because of some unclear cultural reasons, in cases of neurological impairments, men have a higher probability to be taken to a hospital for medical care.

Diverse limitations could be mentioned in the current report. It is based on a particular type of bilingualism; in all the cases Bengali was L1. Bengali, as any language, has certain specific idiosyncrasies, which may potentially impact the brain organization of language. Current results should be replicated using different bilingual samples in order to support the generalizabiliy of our results. Another important limitation refers to the lack of aphasia follow- up. It is known that initial aphasia severity predicts aphasia evolution and recovery (Lazar & Antoniello, 2008). However, the pattern of recovery may be different for L1 and L2 in bilinguals (Paradis, 1977). This is an important type of information that is advisable to include in future studies. An additional limitation refers to the aphasia evaluation; evaluation was carried out in one single language, the L1, in this case Bengali. We do not know the aphasia profile in L2.

It can be anticipated that toward the future, new studies will continue analyzing this critical variable. Its analysis will advance bilingualism understanding, and its effect on the brain organization of language in bilinguals.

## Data Availability

Not applicable

## Notes

### Competing Interest Statement

The authors have declared no competing interest.

